# SARS-CoV-2 detection by rRT-PCR on self-collected anterior nares swabs or saliva compared with clinician-collected nasopharyngeal swabs — Denver and Atlanta, August – November, 2020

**DOI:** 10.1101/2021.02.16.21251521

**Authors:** Grace E. Marx, Sarah E. Smith-Jeffcoat, Brad J. Biggerstaff, Mitsuki Koh, Courtney C. Nawrocki, Emily A. Travanty, Sarah E. Totten, Tracy Scott, Jesse Chavez-Van De Hey, Jesse J. Carlson, Karen A. Wendel, Alexis W. Burakoff, Adam Hoffman, Paulina A. Rebolledo, Marcos C. Schechter, Yun F. Wang, Brooks L. Moore, Hany Y. Atallah, D. Joseph Sexton, Claire Hartloge, Ashley Paulick, Halie K. Miller, Sadia Sleweon, Rebecca Rosetti, Talya Shragai, Kevin O’Laughlin, Rebekah J. Stewart, Juliana da Silva, Caitlin Biedron, CDPHE COVID-19 Laboratory Response Team, CDC COVID-19 Response GA-10 Team, CDC COVID-19 Response Lab Task Force, Jennifer D. Thomas, Hannah L. Kirking, Jacqueline E. Tate, Sarah E. Rowan

## Abstract

Nasopharyngeal swabs (NPS) collected by trained healthcare professionals are the preferred specimen for SARS-CoV-2 testing. Self-collected specimens might decrease patient discomfort, conserve healthcare resources, and be preferred by patients. During August – November 2020, 1,806 adults undergoing SARS-CoV-2 testing in Denver, Colorado and Atlanta, Georgia, provided self-collected anterior nares swabs (ANS) and saliva specimens before NPS collection. Compared to NPS, sensitivity for SARS-CoV-2 detection by rRT-PCR appeared higher for saliva than for ANS (85% versus 80% in Denver; 67% versus 58% in Atlanta) and higher among participants reporting current symptoms (94% and 87% in Denver; 72% and 62% in Atlanta, for saliva and ANS, respectively) than among those reporting no symptoms (29% and 50% in Denver; 50% and 44% in Atlanta, for saliva and ANS, respectively). Compared to ANS, saliva was more challenging to collect and process. Self-collected saliva and ANS are less sensitive than NPS for SARS-CoV-2 detection; however, they offer practical advantages and might be most useful for currently symptomatic patients.

## Introduction

Nasopharyngeal swabs (NPS) are the preferred specimen for SARS-CoV-2 testing by the CDC 2019-nCoV Real-Time Reverse Transcriptase Polymerase Chain Reaction (rRT-PCR) Diagnostic Panel (1). However, NPS collection can be uncomfortable for the patient and should be performed by healthcare professionals in full personal protective equipment (PPE), as the procedure can generate infectious aerosols (2). Prior studies suggest that anterior nares swabs (ANS) and saliva can reliably detect SARS-CoV-2 in patients with coronavirus disease 2019 (COVID-19) (3–5); self-collected ANS or saliva for SARS-CoV-2 detection might decrease patient discomfort, conserve PPE and healthcare resources, and be preferred by patients (6–7).

## Methods

To evaluate self-collected ANS or saliva specimens in real-life SARS-CoV-2 testing settings, participants were enrolled during community testing events in Denver, Colorado, and from clinical settings at a large public hospital in Atlanta, Georgia. After consent, participants were coached to self-collect an ANS and provide a saliva specimen; NPS was then collected by a trained public health professional^*^. Trained interviewers administered structured surveys in English or Spanish to collect demographic characteristics, COVID-19 symptoms^†^, onset of symptoms (Atlanta only), and whether they had close contact^§^ with COVID-19 in the past two weeks; data were entered into Research Electronic Data Capture (REDCap) software (Vanderbilt University). Specimens were handled according to current CDC guidelines^*^ and tested for SARS-CoV-2 using the CDC 2019 nCoV Real-Time RT-PCR Diagnostic Panel assay (1). Results were considered positive when cycle threshold (Ct) values for both 2019 nCoV N1 and N2 PCR targets were <40. Statistical analyses were performed using R software (version 4.0.2; Vienna, Austria) to evaluate and compare test performance of self-collected ANS and saliva specimens with NPS as the reference standard. This activity was reviewed by CDC and was conducted consistent with applicable federal law and CDC policy^¶^. The Institutional Review Boards in Denver (Colorado Department of Public Health and Environment; University of Colorado) and Emory University in Atlanta also determined this activity was an exempt public health activity, consistent with applicable federal law and CDC policy.

## Results

In Denver, 730 participants aged ≥18 years were enrolled at seven SARS-CoV-2 testing events at homeless service sites (n=278) and at 10 community walk-up/drive-through events (n=452). In Atlanta, 1,076 participants aged ≥18 years were enrolled among patients at the emergency department (n=861) or other clinical settings (n=215) at a public hospital. The median age of participants was lower in Denver than in Atlanta (43 versus 52 years); more men than women participated (58% in Denver; 52% in Atlanta) (Table 1). Fewer participants reported Black non-Hispanic race/ethnicity and more participants reported Hispanic ethnicity in Denver than in Atlanta (12% versus 81%; 35% versus 7%). Fewer participants reported current COVID-19 symptoms^†^ but more reported close contact with COVID-19 in Denver than in Atlanta (37% versus 57%; 22% versus 3%).

**TABLE 1.** Characteristics of participants (N=1,806) who contributed saliva, anterior nares swabs, and nasopharyngeal swabs for SARS-CoV-2 detection by rRT-PCR at COVID-19 community testing events and homeless service sites — Denver, CO*, and Atlanta, GA, August – November 2020.

| Characteristic | No. (%) |  |
| --- | --- | --- |
|  | Denver, CO<br>n = 730 | Atlanta, GA<br>n = 1076 |
| **Age group, yrs.** |  |  |
| 18 - 40 | 330 (45.2) | 313 (29.1) |
| 41 - 65 | 334 (45.8) | 619 (57.5) |
| > 65 | 66 (9.0) | 144 (13.4) |
| **Sex** |  |  |
| Male | 420 (57.5) | 559 (52.0) |
| Female | 304 (41.6) | 514 (47.8) |
| Other | 5 (0.7) | 3 (0.2) |
| **Race/Ethnicity** |  |  |
| Black, non-Hispanic | 85 (11.6) | 874 (81.2) |
| White, non-Hispanic | 321 (44.0) | 81 (7.5) |
| Asian, non-Hispanic | 17 (2.3) | 10 (0.9) |
| Hispanic | 257 (35.2) | 71 (6.6) |
| Native American/Alaska Native, non-Hispanic | 14 (1.9) | 2 (0.2) |
| Other, non-Hispanic | 33 (4.5) | 38 (3.6) |
| **Testing location** |  |  |
| Emergency department | N/A | 861 (80.0) |
| Other clinical setting at a public hospital | N/A | 215 (20.0) |
| Community testing event | 452 (61.9) | N/A |
| Homeless service site | 278 (38.1) | N/A |
| **New or worsening symptoms reported at time of testing** |  |  |
| Any symptom | 269 (36.8) | 610 (56.7) |
| Any one of cough, shortness of breath or difficulty breathing, new anosmia or ageusia | 168 (23.0) | 479 (44.5) |
| At least two of fever (measured or subjective) or chills, rigors, myalgia, headache, sore throat, nausea or vomiting, diarrhea, fatigue, congestion/runny nose | 192 (26.3) | 386 (35.9) |
| Flu-like symptoms (Fever AND either cough or sore throat) | 66 (9.0) | 99 (9.2) |
| Fever (measured or subjective) or chills | 90 (12.3) | 147 (13.7) |
| Cough | 79 (10.8) | 324 (30.1) |
| Shortness of breath or difficulty breathing | 86 (11.8) | 346 (32.2) |
| Fatigue | 89 (12.2) | 306 (28.4) |
| Myalgia | 108 (14.8) | 275 (25.6) |
| Headache | 66 (9.0) | 212 (19.7) |
| Anosmia or ageusia | 76 (10.4) | 120 (11.2) |
| Sore throat | 116 (15.9) | 135 (12.6) |
| Congestion or runny nose | 66 (9.0) | 230 (21.4) |
| Nausea or vomiting | 30 (4.1) | 166 (15.4) |
| Diarrhea | 165 (22.6) | 121 (11.2) |
| **Days since symptom onset† (N=590)** |  |  |
| 0–7 | N/A | 420 (71.2) |
| 8–14 | N/A | 84 (14.2) |
| 15–21 | N/A | 30 (5.1) |
| >21 | N/A | 56 (9.5) |
| **Close contact with known COVID-19 case within prior two weeks** | 159 (21.8) | 30 (2.8)<sup>§</sup> |

**Abbreviations:** rRT-PCR = real-time reverse transcriptase polymerase chain reaction; N/A = Not available
\* Participants were enrolled in Denver at seven SARS-CoV-2 testing events at homeless service sites and at 10 community walk-up/drive-through events
† Participants who reported current symptoms at the time of testing with a date of symptom onset (n=20 did not report a date of symptom onset)
§ n=16 additional participants reported close contact but did not report date of exposure and were excluded from this calculation

Of 1,806 total participants, 176 (9.7%) were positive for SARS-CoV-2 by at least one specimen type. Results for both saliva and NPS were available for 496 (67.9%) participants from Denver and 864 (80.3%) from Atlanta; results for valid ANS and NPS were available for 562 (77.0%) participants from Denver and 932 (86.6%) participants from Atlanta. Results for all three specimen types were available for 1,320 (73.1%) participants. Insufficient saliva volume precluded testing in 3% (n=20) and 4.6% (n=49) of participants in Denver and Atlanta, respectively. Using NPS as the reference standard, sensitivity for saliva was 85% and 67% (Table 2) and for ANS was 80% and 58% (Table 3) among participants in Denver and Atlanta, respectively. Sensitivity was higher among participants reporting current COVID-19 symptoms^†^ (94% and 72% for saliva; 87% and 62% for ANS in Denver and Atlanta, respectively) and when saliva and ANS results were combined (87% and 71% in Denver and Atlanta, respectively). Among Atlanta participants, sensitivity was higher among participants reporting more recent symptom onset (80% for saliva; 78% for ANS for onset <7 days; 70% for saliva; 27% for ANS for onset 8–14 days). Among asymptomatic participants, compared to NPS, sensitivity was lower still (29% and 50% for saliva; 50% and 44% for ANS in Denver and Atlanta, respectively). Compared to NPS, negative predictive value was ≥96% for both saliva and ANS among participants at both sites, including among asymptomatic participants. Agreement between saliva and NPS was 0.85 and 0.75 and between ANS and NPS was 0.83 and 0.72 in Denver and Atlanta, respectively (Cohen’s kappa coefficient).

**TABLE 2.** Test performance of saliva compared with clinician-performed nasopharyngeal swabs (NPS) for the detection of SARS-CoV-2 by rRT-PCR, by symptom status at time of testing and by symptom onset — Denver, CO, and Atlanta, GA, August – November 2020.*

|  | Test Characteristic |  |  |  |  |  |  |
| --- | --- | --- | --- | --- | --- | --- | --- |
|  | N | Point Prevalence<br>n (%) | Sensitivity<br>n/N (% [95%CI]) | Specificity<br>n/N (% [95%CI]) | PPV<br>n/N (% [95%CI]) | NPV<br>n/N (% [95%CI]) | Agreement<sup>¶</sup><br>κ (95%CI) |
| **Overall** |  |  |  |  |  |  |  |
| Denver, CO | 496 | 54 (10.9) | 46/54 (85.2 [73.4, 92.3]) | 436/442 (98.6 [97.1, 99.4]) | 46/52 (88.5 [77.0, 94.6]) | 436/444 (98.2 [96.5, 99.1]) | 0.85 (0.77, 0.93) |
| Atlanta, GA | 864 | 64 (7.4) | 43/64 (67.2 [54.3, 78.4]) | 795/800 (99.4 [98.5, 99.8]) | 43/48 (89.6 [77.3, 96.5]) | 795/816 (97.4 [96.1, 98.4]) | 0.75 (0.66, 0.84) |
| **Asymptomatic at specimen collection** |  |  |  |  |  |  |  |
| Denver, CO | 299 | 7 (2.3) | 2/7 (28.5 [8.2, 64.1]) | 289/292 (99.0 [97.0, 99.7]) | 2/5 (40 [11.8, 76.9]) | 289/294 (98.3 [96.1, 99.3]) | 0.32 (0, 0.79) |
| Atlanta, GA | 370 | 14 (3.8) | 7/14 (50.0 [23.0, 77.0]) | 354/356 (99.4 [98.0, 99.9]) | 7/9 (77.8 [40.0, 97.2]) | 354/361 (98.1 [96.0, 99.2]) | 0.6 (0.36, 0.84) |
| **Any symptoms<sup>†</sup> at specimen collection** |  |  |  |  |  |  |  |
| Denver, CO | 197 | 47 (23.9) | 44/47 (93.6 [82.8, 97.8]) | 147/150 (98.0 [94.3, 99.3]) | 44/47 (93.6 [82.8, 97.8]) | 147/150 (98.0 [94.3, 99.3]) | 0.92 (0.85, 0.98) |
| Atlanta, GA | 494 | 50 (10.1) | 36/50 (72.0 [57.5, 83.8]) | 441/444 (99.3 [98.0, 99.9]) | 36/39 (92.3 [79.1, 98.4]) | 441/455 (96.9 [94.9, 98.3]) | 0.79 (0.69, 0.89) |
| **Symptom onset prior to specimen collection (Atlanta, GA only)<sup>§</sup>** |  |  |  |  |  |  |  |
| 0–7 days | 339 | 35 (10.3) | 28/35 (80.0 [63.1, 91.6]) | 302/304 (99.3 [97.6, 99.9]) | 28/30 (93.3 [77.9, 99.2]) | 302/309 (97.7 [95.4, 99.1]) | 0.85 (0.75, 0.94) |
| 8–14 days | 68 | 10 (14.7) | 7/10 (70.0 [34.8, 93.3]) | 58/58 (100 [93.8, 100]) | 7/7 (100 [59.0, 100]) | 58/61 (95.1 [86.3, 99.0]) | 0.8 (0.58, 1) |
| >14 days | 69 | 3 (4.3) | 1/3 (33.3 [0.8, 90.6]) | 65/66 (98.5 [91.8, 100]) | 1/2 (50 [1.3, 98.7]) | 65/67 (97.0 [89.6, 99.6]) | 0.38 (-0.18, 0.93) |

**Abbreviations:** rRT-PCR = real-time reverse transcriptase polymerase chain reaction; PPV = positive predictive value; NPV = negative predictive value
\* Analysis restricted to participants with valid results for both saliva and NP swabs.
† Symptoms defined as new or worsening fever or chills, cough, shortness of breath or difficulty breathing, fatigue, muscle or body aches, headache, new anosmia or ageusia, sore throat, congestion or runny nose, nausea or vomiting, diarrhea.
§ Symptom onset data were collected only from Atlanta participants. n=18 participants reported symptoms but did not specify a date of onset.
¶ Agreement calculated using Cohen’s kappa correlation coefficient statistic.

**Table 3.**
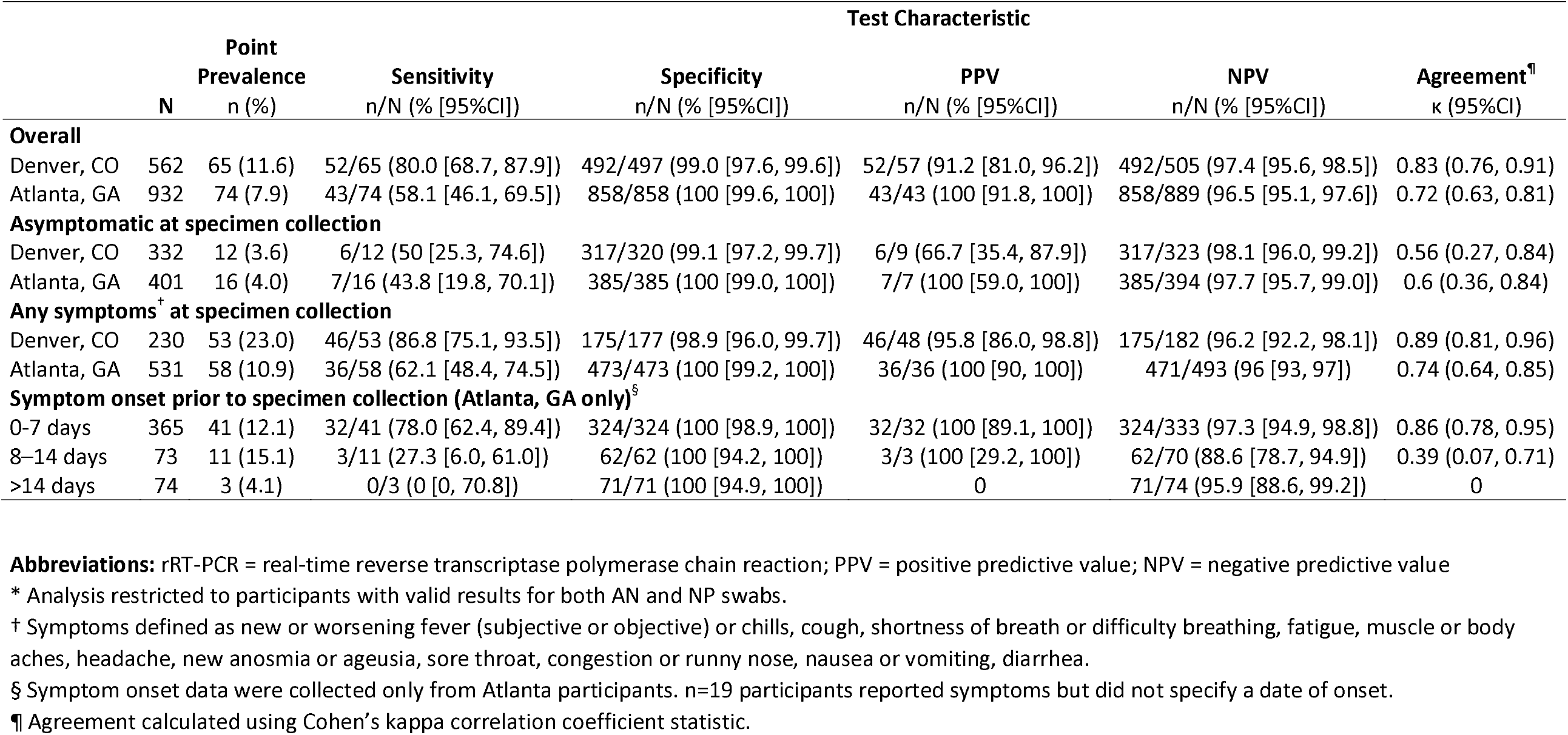
Test performance of self-collected anterior nares swabs (ANS) compared with clinician-performed nasopharyngeal swabs (NPS) for the detection of SARS-CoV-2 by rRT-PCR, by symptom status at time of testing and by symptom onset — Denver, CO, and Atlanta, GA, August – November 2020.*

Ct values from rRT-PCR tests are inversely correlated with the amount of genetic material in the sample (8,9). Studies have shown that SARS-CoV-2 is most often culturable when Ct values are <30 (9,10). Among participants with valid results by all three specimen types and with SARS-CoV-2 detected in NPS, mean NPS Ct values were lowest among participants positive by all three specimen types (18.1 [95% CI 16.9–19.3] and 22.6 [95% CI 20.9–24.3] at the N1 target**; 17.5 [95% CI 16.1–18.8] and 23.1 [95% CI 21.2–24.9] at the N2 target) than among participants positive only by NPS (32.4 [95%CI 25.4–39.4] and 32.9 [95% CI 30.4–35.3] at the N1 target; 32.5 [95% CI 25.1–39.9] and 34.5 [95% CI 31.8 –37.1] at the N2 target) in Denver and Atlanta, respectively. The difference between the mean NPS Ct value among participants positive by only NPS and the mean NPS Ct value among participants positive by all three specimens was 14.3 [95% CI 5.8–22.8] and 10.3 [95% CI 6.9–13.6] at the N1 target, and 15.1 [95% CI 6.0–24.1] and 11.4 [95% CI 7.8–15.0] at the N2 target, in Denver and Atlanta, respectively.

## Discussion

In this investigation, most participants positive for SARS-CoV-2 by rRT-PCR on NPS also tested positive by self-collected saliva and ANS, especially participants reporting current symptoms^†^. Agreement of results between specimen types (saliva and NPS; ANS and NPS) was high for participants reporting current symptoms^†^. Adopting less invasive specimen collection could improve testing uptake, but the benefits of saliva and ANS for testing should be weighed carefully against the loss of sensitivity in asymptomatic people. These findings suggest that several strategies might be applied to optimize SARS-CoV-2 detection in self-collected saliva or ANS. When patients are unable or unwilling to undergo NPS collection, or when PPE or trained healthcare personnel are limited, self-collected saliva or ANS could be offered, with the caveat that some infections might be missed that would have been detected by NPS (13% to 29% of participants in this investigation). Whether cases detected by NPS but negative by saliva/ANS are infectious should be investigated further. Most participants with discordant results between the three specimens had NPS Ct values >30, consistent with decreased genetic material in the sample and potentially non-viable virus *(9,10)*. Self-collected saliva or ANS specimens could be most useful when people report current COVID-19 symptoms^†^, particularly with recent onset. Testing both saliva and ANS in parallel for individual participants appeared to increase sensitivity, but this strategy is an unlikely option; given the increased burden on laboratory resources. Pooled saliva/ANS specimens could be a subject of future research. When developing testing strategies with specific SARS-CoV-2 assays, including non-PCR assays such as antigen-based tests, differences in testing accuracy by specimen type should be considered, along with patient population characteristics.

Sensitivity for both saliva and ANS specimens for the detection of SARS-CoV-2 was higher among participants in Denver than in Atlanta, but the patterns in test performance were the same across sites. The higher test performance among the Denver participants might have been due to differences in healthcare seeking behaviors, testing at different points in the COVID-19 clinical course, or specimen collection, handling, and processing. Participants accessing care in Atlanta received SARS-CoV-2 testing for both clinical concern for COVID-19 and pre-procedure/pre-admission screening; participants in Denver presented to designated SARS-CoV-2 testing events because of concerning symptoms or known/possible close contact.

While saliva specimens had a modestly higher sensitivity than ANS for SARS-CoV-2 detection in this investigation, some participants were unable to produce saliva and others were able to produce saliva in volumes insufficient for testing. Laboratory personnel reported that saliva was difficult to process and often required additional processing to get valid results. The usefulness of saliva as a specimen might be limited if participants are unable to produce adequate specimens or if the laboratory has limited resources to resolve saliva processing issues.

This report has at least five limitations. First, survey responses were self-reported; therefore, responses might be subject to social desirability or recall biases. Second, participants were enrolled in two large urban areas, and might not be representative of other communities. Third, without viral culture, it was not possible to determine if positive results by rRT-PCR represented clinically relevant, infectious virus; however, the higher Ct values among participants testing positive only by NPS (and negative by saliva and ANS) suggests that these individuals might not have had viable virus at specimen collection *(9,10)*. Fourth, slight differences in specimen collection, handling, and processing existed between the study sites in Denver and Atlanta; these may have affected test performance. Fifth, the changing dynamics of SARS-CoV-2 test-seeking behavior and SARS-CoV-2 incidence in Denver and Atlanta during the enrollment period might have affected test performance.

In this investigation, self-collected saliva and ANS specimens were less sensitive for SARS-CoV-2 detection than clinician-obtained NPS; for both specimens, sensitivity was higher among people reporting current COVID-19 symptoms^†^. While use of self-collected saliva and ANS specimens offers practical advantages, challenges to collect and process saliva might be a limitation. Understanding the benefits and limitations of less-invasive specimen collection procedures for SARS-CoV-2 testing should inform public health efforts to design testing programs most appropriate to the local context and population and could possibly improve test uptake.

## Data Availability

All authors had access to the data presented in the manuscript and take responsibility for the analysis and data interpretation.

## Acknowledgments

Participants in Denver, Colorado, and Atlanta, Georgia; Sarah Stella, MD (Denver Health and Hospital Authority); Colorado Coalition for the Homeless; Vyjayanti Kasinathan, MD (Emory University School of Medicine); Luis Lowe (CDC COVID-19 Lab Task; Atlanta, Georgia); Nicolas Wiese (CDC COVID-19 Lab Task Force; Atlanta, Georgia)

## Disclaimer

The findings and conclusions in this report are those of the authors and do not necessarily represent the official position of the U. S. Centers for Disease Control and Prevention (CDC).

## Summary

### What is already known about this topic?

Limited data are available on the comparative usefulness of self-collected ANS and saliva for SARS-CoV-2 detection, compared with NPS in real-world settings.

### What is added by this report?

Sensitivity of saliva and ANS for SARS-CoV-2 detection was lower than NPS. Compared with NPS, saliva and ANS sensitivity appeared higher among people reporting symptoms, but <50% for saliva and ANS among participants reporting no symptoms.

### What are the implications for public health practice?

Understanding practical advantages and limitations to self-collected ANS and saliva specimens for SARS-CoV-2 detection can inform testing programs.

## Footnotes

* https://www.cdc.gov/coronavirus/2019-ncov/lab/guidelines-clinical-specimens.html

† COVID-19 symptoms in this investigation were defined as reports of new or worsening fever (measured or subjective) or chills, cough, shortness of breath or difficulty breathing, fatigue, myalgia, headache, anosmia or ageusia, sore throat, congestion or runny nose, nausea or vomiting, or diarrhea.

§ https://www.cdc.gov/coronavirus/2019-ncov/php/contact-tracing/contact-tracing-plan/appendix.html#contact

¶ This activity was reviewed by CDC and was conducted consistent with applicable federal law and CDC policy: 45 C.F.R. part 46.102(l)(2), 21 C.F.R. part 56; 42 U.S.C. Sect. 241(d); 5 U.S.C. Sect. 552a; 44 U.S.C. Sect. 3501 et seq.

** https://www.cdc.gov/coronavirus/2019-ncov/lab/virus-requests.html

